# Identifying facilitators and barriers to culturally responsive communication for racial, ethnic, sexual, and gender minoritized patients when screened for COVID-19 vaccinations: A Scoping Review Protocol

**DOI:** 10.1101/2023.08.11.23293975

**Authors:** Nikhil Kalita, Patrick Corr, Maranda C. Ward, Julia Xavier, Paige McDonald

**Author notes:** Corresponding Author, (NK); 2600 Virginia Avenue NW, Suite 300, Washington DC, 20037. These authors contributed equally to this work and have approved the following manuscript for submission to PLOS ONE.

## Abstract

**Introduction:** Racial, ethnic, sexual, and gender minoritized groups are considered historically excluded groups and have been disproportionately affected by the coronavirus disease 2019 (COVID-19) pandemic. The influence of social determinants of health (SDOH), including access to screening and treatment, and other systemic and structural factors are largely responsible for these disparities. Primary care practitioner (PCP) competence in culturally responsive screening practices will be critical to reducing the impact of systemic and structural factors serving as barriers to screening and treatment. Correspondingly, improving the capacity of PCPs to communicate with patients in a culturally responsive manner may influence improved screening and treatment outcomes for minoritized groups related to COVID-19. This scoping literature review aims to determine the current breadth of literature on culturally responsive communication (CRC) in regard to COVID-19 vaccination screening for historically excluded, or minoritized groups. Results from this review will inform the development of a training series and social marketing campaign to improve PCPs capacity in CRC.

**Objectives:** This scoping literature review aims to analyze existing literature on culturally responsive COVID-19 vaccinations between PCPs and patients in the U.S., specifically for racial, ethnic, sexual, and gender minoritized groups. Results of this scoping review will inform the development of a training series and social marketing campaign to improve capacity of PCPs in this area. Additionally, the review will inform recommendations for future research.

**Materials and Methods:** This scoping review will be performed following the framework of Arksey and O’Malley and the Preferred Reporting Items for Systematic Reviews and Meta-Analyses extension for scoping reviews (PRISMA-ScR). Relevant studies between the years 2019-2022 were identified using a rigorous search strategy across four databases: MEDLINE (via PubMed), Scopus, Cochrane (CENTRAL; via Wiley), and CINAHL (via EBSCO), using Boolean and Medical Subject Headings (MeSH) search terms. Studies will be uploaded to the data extraction tool, Covidence, to remove duplicates and perform a title/abstract screening, followed by a full-text screening.

**Results:** The data extraction and analysis phases of the scoping review are in progress. Data will be analyzed for themes related to culturally responsive COVID-19 screening practices in clinical encounters with the identified study populations. Results will be reported by theme and align to PRISMA-ScR guidelines.

**Discussion:** To our knowledge, this is the first study to use scoping methods to investigate the barriers and facilitators to CRC of COVID-19 vaccine screening for historically excluded communities in the U.S. The work and results from this research will be directly utilized for the development of nationally-accessible, continuing medical education materials to teach PCPs about CRC, as well as other materials to influence relevant policy changes within the healthcare landscape.

## Introduction

The impact of racism, heterosexism, and transphobia in healthcare settings has been elucidated by the coronavirus disease 2019 (COVID-19) pandemic. People from historically excluded communities, such as those who are racial, ethnic, sexual, and gender minoritized, endure disproportionate systemic barriers and structural inequities related to social determinants that influence increased susceptibility to disease and associated health outcomes.^1–3^ For example, both COVID-19 and human immunodeficiency virus (HIV) are both preventable, communicable viruses that are highly stigmatized and disproportionately affect health outcomes for these minoritized groups.^4–5^ Culturally responsive communication in the primary care setting for COVID-19 and HIV among these groups may mitigate the negative effects of structural and systemic barriers to care. To promote simultaneous culturally responsive screening for COVID-19 and HIV, our study team aims to design training modules to build PCP capacity in culturally responsive communication (CRC). We are conducting two separate scoping reviews to inform and validate the design of this training, corresponding social marketing campaigns, and policy recommendations. The protocol for one scoping review related to culturally responsive communication for HIV and PrEP has been published^6^ with its analysis underway.

This protocol informs a scoping review focused on understanding how, and whether CRC is occurring between PCPs and minoritized groups related to COVID-19 vaccination screening. As noted in the aforementioned published protocol, the data from the two reviews will be combined to inform 1) a training series for PCPs pertaining to CRC screening for COVID-19, HIV testing and PrEP screening, 2) a social marketing campaign by PCPs to encourage other PCPs to routinize culturally responsive conversations about testing, screening, and prevention; and 3) a white paper with policy recommendations for improved screening guidelines for HIV and to inform better implementation of current guidelines for HIV, PrEP/PEP and COVID-19.^6^ The remainder of this article describes the protocol guiding the scoping review on CRC related to COVID-19 screening by PCPs with the noted minoritized groups.

As there are many intersecting themes between the disproportionate burden of COVID-19 endured by racial, ethnic, sexual, and gender minoritized patients, PCPs should learn to acknowledge the potential differences in culture and perspective when communicating with their patients during the screening of COVID-19 vaccines. This requires PCPs to gain an appreciation for CRC and learn how to facilitate it. However, PCPs may not have the appropriate skills and training to engage in non-judgmental conversations with historically excluded communities about various aspects of care.^7–11^

In existing published literature, CRC has been related to culturally competent care and has been defined as “communicating with awareness and knowledge of cultural differences and attempting to accommodate those differences,”^12(p2)^ and it necessarily involves “respect and an understanding that sociocultural issues such as race, gender, sexual orientation, disability, social class and status can affect health beliefs and behaviours”.^12(p2)^ Xavier & colleagues^6^ expand upon this definition, while emphasizing that within primary care, PCPs must “engage with patients” in a responsive way that appreciates the role of culture, including their views as healthcare professionals. According to these authors:

> “Cultural responsiveness centers unique patient experiences and understandings of health and illness, recognizes the individual biases that clinicians may hold, and seeks to work productively with patients who are not typically represented or valued in the Western understandings of care. At an organizational level, cultural responsiveness includes valuing diversity within the community; institutionalizing cultural awareness; and adapting to best serve the community by creating policies, systems, administrations, and protocols that allow for effective cross-cultural interactions. This type of approach allows healthcare practitioners to work consciously and effectively toward cultivating health equity for historically marginalized groups.^6(p2)^”

Correspondingly, CRC could be instrumental in addressing the burden of COVID-19 in historically excluded groups, if we had greater understanding of if and how CRC occurs in patient-practitioner interactions, particularly with minoritized populations. Additionally, current outcome assessments only relate to the influence of cultural competence training on the PCP^13^ and on few patient outcomes, such as satisfaction and compliance.^14^ Additional knowledge is required to understand the process, PCP training outcomes, and patient outcomes regarding CRC.

The study team aims to build the capacity of PCPs to routinize CRC in COVID-19 vaccination screening visits. As such, this scoping review focuses on investigating what has been published on CRC between PCPs and historically excluded populations related to COVID-19 vaccination screening.

## Background

### Disproportionate COVID-19 Outcomes

Racial and ethnic minoritized communities are at an increased risk of exposure and burden from COVID-19 due to many systemic disadvantages. COVID-19 disproportionally affects historically excluded communities due to a lack of access to healthcare, racism, gender oppression, structural discrimination, medical mistrust, and more.^15–16^ Racial and ethnic minoritized patients have about one and a half times greater risk of COVID-19 infection and are twice as likely to die from COVID-19 as their white counterparts when accounting for age differences across racial and ethnic groups.^17^ In fact, in the summer of 2020, Hispanic people were five times more likely to die from COVID-19, and Black people were three times as likely to die from COVID-19 compared to their white counterparts.^17^ Endemic inequities are also persistent when it involves income, education, nutrition, transportation, housing, jobs, environment, psychosocial stress, and health care.^18^ Each of these inequities can be directly tied to the disproportionate incidence, burden, and mortality of COVID-19 for racial and ethnic minoritized people.^18^ For example, Black Americans are exposed to air that is 38% more polluted compared to white Americans, increasing their propensity for developing asthma as well as their subsequent risk of COVID-19.^19^

Similar to racial and ethnic minoritized groups, sexual and gender minoritized groups face systemic disparities in relation to COVID-19. There is little known on the overall health effects of COVID-19 for sexual and gender minoritized groups due to the limited data collection and reporting executed by the U.S. public health system.^20^ Despite this, U.S. sexual and gender minoritized people reported having significant and disproportionate poor mental health outcomes due to the COVID-19 pandemic.^21–25^ Additionally, sexual and gender minoritized patients reported greater rates of job loss, housing, and food insecurity along with minority stress and stigmatization, all linked to higher levels of mental illness during the COVID-19 pandemic.^21–27^

### COVID-19 Vaccination Acceptance

Disparities in vaccine access and acceptance are also associated with disproportionate COVID-19 hospitalization and mortality among racial, ethnic, sexual, and gender minoritized patients. By the end of April, 2021, 47% of Hispanic Americans and 46% of Black Americans had received at least one vaccine, compared to 59% of white Americans.^28^ As of July 6, 2021, only 44% and 41% of Black and Hispanic Americans received a COVID-19 booster dose, while 56% of White Americans received a COVID-19 booster dose.^29^ Many recent equity-based efforts may have caused vaccine uptake to increase among all racial and ethnic minoritized groups.^29^ Though vaccine uptake by all groups has recently equalized, it is important to understand the influences on initial disproportionate uptake by racial and ethnic minoritized populations. Additionally, sexual and gender minoritized patients faced initial barriers to receiving and accepting COVID-19 vaccinations with many intersecting themes when compared to racial and ethnic minoritized patients.^27^ A persistent barrier for COVID vaccinations among sexual and gender minoritized groups include historical and ongoing medical trauma. These barriers, faced by all historically excluded groups, should be addressed in primary care settings.

COVID-19 vaccination acceptance is directly related to psychological behavior, societal and political issues, and vaccine-derived factors that strongly influence decision-making.^30^ Vaccine-related behavioral patterns are complex and influenced by various extrinsic factors.^30^ Negative extrinsic factors can accumulate and prevent people from receiving vaccines.^31^ A major negative extrinsic factor is the behavior of medical mistrust significantly derived from the malicious history of the mistreatment of historically excluded populations in healthcare and medical research.^32^ In the 1800s, James Marion Sims performed nonconsensual, experimental surgeries on several enslaved Black women without anesthesia.^33^ From 1932 to 1972, 600 Black men with syphilis were examined without proper consent nor access to penicillin treatments that were readily available.^15^ In the late 1900s, Black women in Mississippi, who had gone to receive surgeries for their benign tumors, instead had their uterus removed without their consent.^15^ Doctors in the 1980s falsely referred to AIDS as “Gay-Related Immune Deficiency,” kick-starting the stigmatization of HIV and AIDS against sexual and gender minoritized patients.^34^ These profound historical mistreatments have partly caused the justified mistrust in healthcare among minoritized populations. Medical mistrust could also arise from present-day extrinsic factors that include communication content, communication presentation, policy, and vaccine delivery in clinical settings.^35^

### COVID-19 Vaccination Communication and Culturally Responsive Communication

COVID-19 demonstrated the importance of CRC in primary care settings during an emergent health crisis. Throughout COVID-19, the general public relied on varying sources of information to determine COVID-19 vaccine safety and efficacy, some of which are not reliable or evidence-informed.^36^ This reliance, highlights the central role of PCPs as reputable sources of evidence-informed guidance for their patient populations. PCPs are often responsible for improving health literacy by communicating evidence-based, understandable, and accessible health information to patients.^37^ Unfortunately, disparate treatment and communication between PCPs and their minoritized patients exists.^38^ Racial, ethnic, sexual, and gender minoritized patients are significantly more likely to report discrimination, a notable predictor of medical mistrust.^15, 39^ Communication disparities, specifically, are linked to PCP bias and stigma, leading to increased mistrust of healthcare practitioners along with other barriers significantly affecting patient adherence and healthcare-seeking behavior.^38^ PCPs can better address these current disparities in the clinical setting through our expanded definition of CRC.^12^ However, we first need to learn more about if and how CRC is currently occurring in PCP encounters with minoritized patients.

## Objectives

A scoping review rapidly maps the body of literature on a specific research area and the main sources and types of evidence available.^40^ Scoping reviews can develop a basis or preliminary understanding of published literature on a topic before conducting systematic reviews.^41^ This scoping review aims to summarize and disseminate information on CRC between PCPs and racially, ethnically, sexually, and gender minoritized patients related to COVID-19 vaccination. The results of this scoping review will be used to inform future research and policy recommendations to understand and improve PCPs’ capacity to routinize COVID-19 screening and prevention with all patients and rely on CRC for patients from historically excluded communities.

## Materials and Methods

This scoping review will be conducted in accordance with the Arksey and O’Malley methodological framework.^40^ The framework provides a flexible design for when researchers redefine search terms as familiarity with the literature increases.^40^ The process is considered to be iterative for researchers to engage with each stage, so that the review is fully comprehensive.^40^ The framework suggests a scoping review undergo five stages: (1) identify the research question; (2) identify relevant studies; (3) select studies and extract data; (4) chart the data; and (5) collate, summarize, and report results.^40^ This scoping review will also be guided by the specific steps of the PRISMA extension for scoping reviews (PRISMA-ScR).^42^

### Institutional review board statement

This project did not utilize human subjects, nor did it involve a process of informed consent as the need for consent was waived by an ethics committee. This manuscript exclusively provides an overview of a scoping review protocol. The data from this scoping review will be used to inform a continuing medical education intervention that is IRB-approved and supported by grant funding.

### Stage 1: Identifying the Research Question

Before proceeding with the scoping review, the research team first identified pertinent issues while crafting an initial research question to bring forward to multiple clinicians for review and feedback. Most PCPs consulted expressed that there should be a focus on understanding and teaching CRC regarding COVID-19 vaccinations rather than on general COVID-19 prevention and screening. As of now, COVID-19 vaccinations are still not widely accepted in the U.S. despite the compounding positive effects of herd immunity.^43^ Additionally, minoritized populations are disproportionately affected by COVID-19 due to various systemic and interpersonal barriers described previously. Therefore, the concern for appropriate culturally tailored interventions with COVID-19 vaccination is particularly valid. Correspondingly, the research question (PRISMA-ScR Item 4: Objectives) guiding this review is: “*How is culturally responsive communication occurring between patient and practitioner related to COVID-19 vaccination and booster screening for racially, ethnic, sexually, and gender minoritized patients?”*

### Stage 2: Identifying Relevant Studies

The search was conducted across the four databases (PRISMA-ScR Item 7: Information Sources) of MEDLINE (Pubmed), Scopus, CENTRAL (Cochrane Central Registry of Controlled Trials), and CINAHL (Complete). Studies with various title-abstract and Boolean and Medical Subject Headings (MeSH) terms pertaining to the research question and definitions of key concepts were included in the search strategy. With input from the research team, collaborators, and an experienced research librarian, an initial search strategy was devised (see Table 1). The search strategy required four categories of terms: terms including the population of interest, terms similar to “culturally competent”, terms synonymous to “COVID-19”, and terms related to vaccination in the context of the study.

**Table 1:** MEDLINE Search Strategy.

| Categories | Search Terms |
| --- | --- |
| Population of Interest | (Marginaliz* [tiab] OR disadvantag* [tiab] OR underserv* [tiab] OR vulnerable populations [mesh] OR medically underserved area [mesh] OR LGBT* [tiab] OR BIPOC [tiab] OR POC [tiab] OR minorit* [tiab] OR ethnic and racial minorities [mesh] OR minority groups [mesh] OR minority health [mesh] OR ethnic minorit* [tiab] OR racial minorit* [tiab]<br>OR gay [tiab] OR lesbian* [tiab] OR homosexual* [tiab] OR health services for transgender persons [mesh] OR sexual and gender minorities [mesh] OR sexual minorit* [tiab] OR gender minorit* [tiab] OR homosexuality [mesh] OR homosex* [tiab] OR transgender* [tiab] OR transgender persons [mesh] OR transsexualism [mesh] OR transex* [tiab] OR MSM [tiab] OR WSW [tiab] OR YMSM [tiab] OR men who have sex with men [tiab] OR bisexual* [tiab] OR queer* [tiab] OR nonbinary [tiab] OR intersex [tiab]<br>OR indigenous* [tiab] OR American Native Continental Ancestry Group [mesh] OR health services, indigenous [mesh] OR indigenous peoples [mesh] OR alaskan native* [tiab] OR indigenous canadians [mesh] OR native american* [tiab] OR native-american* [tiab] OR native* [tiab] OR nation people* [tiab] OR inuit* [tiab] OR inuits [mesh] OR indian* [tiab]<br>OR African American* [tiab] OR POC [tiab] OR people of color [tiab] OR African-american* [tiab] OR black* [tiab] OR blacks [mesh] OR African Americans [mesh] OR health disparity, minority and vulnerable populations [mesh]<br>OR hispanic [tiab] or latino [mesh] OR hispanic* [tiab] OR latino* [tiab] OR latinX* [tiab] OR latina* [tiab]<br>OR asian americans [mesh] OR asian* [tiab]<br>OR pacific islander americans [mesh] OR native hawaiian [mesh]<br>OR prejudice [mesh] OR bias [tiab] OR prejudice [tiab] OR racis* [tiab] OR racia* [tiab] OR sexis* [tiab] OR discriminat* [tiab] OR homophob* [tiab] OR inequit* [tiab] OR inequalit* [tiab] OR health inequities [mesh] OR healthcare disparities [mesh] OR disparit* [tiab] OR social inequalit* [tiab] OR racial inequalit* [tiab] OR segregat* [tiab] OR social stigma [mesh] OR stereotyping |
|  | [mesh] OR social discrimination [mesh] OR social marginalization [mesh] OR social isolation [mesh] OR stigma* [tiab] )<br>**AND** |
| Culturally Competent | ( cultural* responsi* [tiab] OR culturally-responsive [tiab] OR culturally competent care [mesh] OR cultural* competen* [tiab] OR culturally-competent [tiab] OR cultural* aware* [tiab] OR culturally-aware [tiab] OR cultural* sensitiv* [tiab] OR culturally-sensitive [tiab] OR cultural* congruen* [tiab] OR culturally-congruent [tiab] OR cross-cultur* [tiab] OR cross cultur* [tiab] OR cultural* grounded* [tiab] OR culturally-grounded [tiab] OR inclusi* [tiab] OR competen* [tiab] OR cultural* adapt* [tiab] OR culturally-adapted [tiab] OR cultural* tailor* [tiab] OR culturally-tailored [tiab] OR culturally influenced [tiab] OR culturally-influenced [tiab] OR affirm* [tiab] OR transcultural [tiab] OR multicultural [tiab] OR intercultural [tiab] OR cultural* litera* [tiab] OR cultural* respect* [tiab] OR cultural* appropriate* [tiab] OR culturally accept* [tiab] OR cultural* safe* [tiab] OR cultural* intelligen* [tiab] OR<br>patient communic* [tiab] OR patient interact* [tiab] OR patient satisfact* [tiab] OR patient relation* [tiab] OR patient trust* [tiab] OR patient concordanc* [tiab] OR patient trust* [tiab] OR patient collaborat* [tiab] OR patient partner* [tiab] OR patient-centered* [tiab] OR patient centered* [tiab] OR patient orientat* [tiab] OR patient-orientat* [tiab] OR Patient-Practitioner Orientation Scale [tiab] OR PPOS [tiab] OR provider communic* [tiab] OR provider interact* [tiab] OR provider satisfact* [tiab] OR provider relation* [tiab] OR provider trust* [tiab] OR provider concordanc* [tiab] OR provider trust* [tiab] OR provider collaborat* [tiab] OR provider partner* [tiab] OR patient-centered* [tiab] OR patient centered* [tiab] OR Patient-Practitioner Orientation Scale [tiab] OR PPOS [tiab] OR Health Communication [mesh] OR Patient Satisfaction [mesh] OR Health Education [mesh] OR patient-practitioner* [tiab] OR patient-provider* [tiab] OR patient-physician* [tiab] OR patient-doctor* [tiab] OR practitioner-patient* [tiab] OR provider-patient* [tiab] OR physician-patient* [tiab] OR doctor-patient* [tiab] )<br>**AND** |
| COVID-19 | (COVID-19 [mesh] OR COVID* [tiab] OR coronavirus* [tiab] OR corona virus* [tiab] OR SARS-CoV-2 [mesh] OR SARS-CoV-2* [tiab] OR COVID-19* [tiab] OR nCoV* [tiab] OR SARS Coronavirus 2* [tiab] OR SARS Corona virus 2* [tiab] OR Severe Acute Respiratory Syndrome Coronavirus 2* [tiab] OR Severe Acute Respiratory Syndrome Coronavirus 2* [tiab] OR Severe Acute Respiratory Syndrome Coronavirus-2* [tiab])<br>**AND** |
| Vaccination | (vaccin* [tiab] OR Vaccines [mesh] OR booster* [tiab] OR immuniz* [tiab]) |

Several groups within historically excluded populations are named in different contexts, languages, and forms. So, it was crucial to include all possible terms that each marginalized group is referred to in scientific literature. For example, each term and all synonyms within the acronyms of LGBTQIA+ and BIPOC were searched. Terms that potentially cause marginalization for groups such as “discrimination”, “prejudice”, “stereotyping”, were also included. All terms related to cultural responsiveness and the practitioner-patient relationship, such as “cultural competence”, “cultural sensitivity”, and “patient-centered” were included.

As COVID-19 has surged into the global environment, various nomenclature of COVID-19 have also emerged. Studies may have different terminology of COVID-19 due to their scientific nature and specificity of results. Therefore, a thorough identification process of all possible COVID-19 terms was implemented in the search strategy. The differing names include, but were not limited to, “COVID”, “n-CoV2”, “SARS Coronavirus 2”. Search terms specifically regarding vaccination were the last set of terms that had to be included in the search. These terms refer to primary prevention and include “vaccination”, “booster”, “immunization”.

The finalized search strategies brought about the following results by database: MEDLINE yielded 284 results; SCOPUS yielded 545 results; CENTRAL yielded 61 reviews and 114 prospective clinical trials; and CINAHL database yielded 127 results. All databases posed problems with formatting and character technicalities. Correspondingly, a few adjustments were made to conduct the search appropriately and reflect searches in other databases. Table 1 provides an overview of the MEDLINE search strategy (PRISMA-ScR Item 8: Search), translated and utilized in the other databases.

### Stage 3: Study Selection

The scoping review will incorporate two levels of screening using Covidence literature review software. First, titles and abstracts will be reviewed for all manuscripts, and consensus will be required from at least two reviewers for inclusion. All studies deemed relevant in the title and abstract review shall move forward for review in the full-text level of screening (PRISMA-ScR Item 9: Selection of Sources of Evidence).

In the title and abstract review, four investigators (NK, JX, SP, DB) will independently screen each study based on the following inclusion criteria:

- Study must be based in the U.S. or analyze a U.S. population (unless it is a scoping or systematic review)
- Study must conduct research for or with an LGBTQIA+ and/or BIPOC group
- Study must include any results and/or discussion related to COVID-19 vaccines
- Study must include attitudes, behaviors, etc. of patients and/or PCPs in healthcare settings
- Study must have been published after November 2019 (start of COVID-19)
- Study must not be a protocol or any type of research not already published

The search will also be limited (PRISMA-ScR Item 6: Eligibility Criteria) to studies that concern COVID-19 vaccination screening for minoritized populations. Studies will be included if both investigators found that they fulfilled all the requirements of the inclusion criteria. If two investigators have differing opinions on a study, a third investigator will make a final decision on the study’s inclusion. The title and abstract review will be conducted by primary reviewers (NK, JX) and secondary reviewers (SP, DB).

Full-text screening of the studies included from the title and abstract review will then be conducted. This stage will require two reviewers to read articles in their entirety and decide whether they should be included in the review. Similar to the title and abstract screening, if two investigators have differing opinions on a study, a third investigator decides if it should be included in the review to be forwarded for data extraction. The relevance and inclusion criteria of the full-text screening will be the same as that of the title and abstract screening. The full-text screening will be conducted by primary reviewers (NK, JX) and a new team of secondary reviewers (SP, PC). The resulting studies will qualify for inclusion in the next step of the review, the extraction phase, where data on these studies will be charted.

### Stage 4: Charting the Data

This stage is meant to collate and synthesize the data in a comprehensive and organized manner to appropriately extract information relevant to our research question (PRISMA-ScR Item 10: Data Charting Process). The extraction phase will be moved from Covidence to Google Sheets to allow for better collaboration and cohesiveness. This study team deemed that Covidence has several limitations when collaborating with group members, copying and pasting from full-text presentations of studies, and selection capabilities. Google Sheets will also allow more flexibility for complicated questions and connectivity between team members. Information from the included studies will be reviewed by 9 reviewers (NK, PM, PC, AK, MW, HC, PS, OC, MCW) and extracted through an evidence-based format of prompts and questions requiring a specific input, or checkbox selection, through Google Sheets (PRISMA-ScR Item 11: Data Items).

**Table 2:** Format of prompts and questions for extraction phase.

| **Prompt/Question (s)** | **Input; Checkbox Selection** |
| --- | --- |
| Reviewers | Reviewers' initials. |
| Covidence ID | ID provided by Covidence. |
| Author(s) | Name of first author. |
| Publication Title | Title of study. |
| Publication Year | Year when study was published. |
| Study Selection | Location where study was conducted. |
| Study Design | Checkbox Selection:<br>• Randomized controlled trial<br>• Non-randomized experiment<br>• Cohort study<br>• Cross-sectional study<br>• Quantitative research<br>• Participatory Action Research<br>• Systematic review<br>• Case series<br>• Case report<br>• Diagnostic test accuracy study<br>• Opinion Piece/editorial<br>• Other (*space to input design provided*) |
| Intervention (if applicable) | Checkbox Selection:<br>• Quality improvement (i.e. protocol, screening QI)<br>• Community health/Public health initiatives<br>• Patient education: unspecified<br>• Patient education: knowledge/attitudes<br>• Patient education: skill-building<br>• Patient education: behavior change<br>• Clinical education: unspecified<br>• Clinician education: knowledge/attitudes<br>• Clinician education: skill-building (i.e. measurable clinical tests/skills)<br>• Clinician education: CME courses<br>• N/A<br>• Other (*space to input intervention type provided*) |
| Timeframe of Study | Timeframe of data collection of study. |
| Study Aims | Verbatim copy and paste of aims indicated by study. |
| Study Population | Specific population(s)/group(s) studied, including any demographic details |
| Methodology Overview | Checkbox Selection:<br>• Observation/Participant observation<br>• Literature review (i.e. systematic review, scoping reviews, etc.)<br>• Art-based forms (i.e. photo, voice data poems)<br>• Surveys/Questionnaire<br>• Individual Interviews<br>• Paired Interviews<br>• Focus groups<br>• Administering intervention and tracking outcomes<br>• Biometric data (i.e. fitbits and cardiac health)<br>• Secondary data analysis/archival study (i.e. hospital-based, EMR, etc.)<br>• Other (*space to input methodology overview provided*) |
| Results | All results copied and pasted. |
| Level of Communication Addressed | Checkbox Selection:<br>• Patient-practitioner interaction<br>• EHR/patient portal communication<br>• Public health communication - local level (i.e. local non-profits, community messages)<br>• Public health communication - national level (i.e. government orgs, large orgs)<br>• Social media<br>• N/A<br>• Other (*space to input level of communication addressed*) |
| How does it address racial and ethnic minoritized patients? | Checkbox Selection:<br>• Engaged racial and ethnic minoritized groups as study participants<br>• Offered recommendations specific to needs of racial and ethnic minoritized patients<br>• Collaborated with racial and ethnic minoritized groups or related organizations<br>• Racial and ethnic minoritized people participated in study analysis<br>• Racial and ethnic minoritized people leading/designing study<br>• N/A |
|  | Other (*space to input the addressing of racial and ethnic minoritized patients*) |
| How does it address sexual and gender minoritized patients? | Checkbox Selection:<br>Engaged sexual and gender minoritized groups as study participants<br>Offered recommendations specific to needs of sexual and gender minoritized patients<br>Collaborated with sexual and gender minoritized groups or related organizations<br>Sexual and gender minoritized people participated in study analysis<br>Sexual and gender minoritized people leading/designing study<br>N/A<br>Other (*space to input the addressing of sexual and gender minoritized patients*) |
| How does it address COVID-19 Vaccine Communication? [EXPLANATION] | Reviewers explain why they indicated the specific component as a facilitator or barrier to COVID-19 vaccine communication addressed by the study. |
| How does it address COVID-19 Vaccine Communication? [QUOTES] | Reviewers input excerpts of the study supporting their choice of indicating the specific component as a facilitator or barrier to COVID-19 vaccine communication addressed by the study. |
| Implications | Verbatim or summarized input of implications, recommendations, etc. explained by authors of the study. |
| Limitations | Verbatim copied and pasted of limitations or biases indicated by authors of the study. |

To increase the rigor of the scoping literature review, 3 primary reviewers (NK, PM, PC) will undergo another review of all 81 studies to determine if there was substantial mention of CRC, specifically in regard to direct PCP communication with patients. The 3 reviewers (NK, PM, PC) will first extract data on CRC between PCPs and patients from all 81 studies through a similar process as the initial data extraction utilizing Google Sheets. If this information was present in a substantial amount, or in at least one sentence throughout the individual studies, they will be included for data analysis. All results from these reviews will be conducted individually by the 3 reviewers (NK, PM, PC) and then be reviewed amongst each reviewer to finalize decisions on what data should be analyzed and what should not.

### Stage 5: Collating, Summarizing, and Reporting the Results

Data determined to be useful in answering the study question, “How is CRC occurring between patient and practitioner related to COVID-19 vaccination and booster screening for racially, ethnically, sexually, and gender minoritized patients?” will be collated again via Google Sheets. The data will then be organized by theme and relevance to determine the scope of literature regarding our topic of interest, along with potential gaps in existing literature. The themes and relevance will be identified inductively through emergent coding and then deductively through the lenses of Critical Race Theory,^44^ Queer Theory,^45^ and the Socio-Ecological Framework.^46^ These perspectives stress the importance of centering the voices of minoritized patients in order to understand, disrupt, and reshape systems of power, as these groups are best able to speak to their humanity and experiences. The scoping literature review will also focus on identifying the breadth of the available literature rather than its quality, which is typically evaluated through a systematic literature review.^41^ After analysis, results will be synthesized and reported according to PRISMA-ScR guidelines (Item 13: Synthesis of Results; Items 15-19). The process used to select studies will be detailed in a PRISMA flowchart (PRISMA-ScR Item 14: Selection of Sources of Evidence). The study team will disseminate our findings of the scope of available literature, as well as opportunities for future research and clinical interventions in regard to CRC for COVID-19 vaccination screening in primary care settings.

## Discussion

As minoritized patients often face many barriers to health care, PCPs must aim to be trusted sources of information through utilizing CRC to facilitate important discussions with minoritized patients about COVID-19 vaccinations. Discussions incorporating CRC are vital in influencing a patient’s decision-making process when considering to take a COVID-19 vaccination or booster. This scoping review will indicate if and how CRC is currently implemented in PCP encounters with minoritized patients. To our knowledge, this is the first study to use scoping methods to investigate the barriers and facilitators to CRC between PCPs and minoritized patients regarding COVID-19 vaccine screening. This scoping review protocol will allow us to adequately map the landscape, gaps, and prominent themes of current research. Our findings will then be disseminated in publication and via nationally-accessible, continuing medical education materials, as well as other materials to influence relevant policy changes within the healthcare landscape. One limitation involved the translation of search terms of interest across databases in the health and medical sciences. Each database has unique language parameters and search requirements, resulting in minor differences across databases.

## Supporting Information

PRISMA-P 2015 checklist.docx

## Acknowledgments

The authors would like to acknowledge Thomas Harrod, Associate Director of Reference, Instruction, and Access at the George Washington University’s Himmelfarb Health Sciences Library for his guidance and support in developing the search strategies for this scoping review. The authors would like to acknowledge Saylor Pershing, Darrell Bailey, Hasina Chimeka-Tisdale, Olivia Cristillo, Paloma Delgado, Madeleine Will, and Abigail Konopasky for their assistance in the text screening and extraction process.

## Data Availability

All relevant data from this study will be made accessible upon completion of the scoping review. No datasets were produced or analyzed in the current study.

## References

1. Abedi V, Olulana O, Avula V, et al. Racial, Economic, and Health Inequality and COVID-19 Infection in the United States. J Racial Ethn Health Disparities. 2021;8(3):732–742. doi:10.1007/s40615-020-00833-4

2. Sears B, Conron KJ, Flores AR. The Impact of the Fall 2020 COVID-19 Surge on LGBT Adults in the US. University of California, Los Angeles. February 2021. Accessed May 24, 2023. https://williamsinstitute.law.ucla.edu/publications/covid-surge-lgbt/.

3. Yancy CW. COVID-19 and African Americans. JAMA. 2020;323(19):1891–1892. doi:10.1001/jama.2020.6548

4. Rewerska-Juśko M, Rejdak K. Social Stigma of Patients Suffering from COVID-19: Challenges for Health Care System. Healthcare (Basel). 2022;10(2):292. doi:10.3390/healthcare10020292

5. Rueda S, Mitra S, Chen S, et al. Examining the associations between HIV-related stigma and health outcomes in people living with HIV/AIDS: a series of meta-analyses. BMJ Open. 2016;6(7):e011453. doi:10.1136/bmjopen-2016-011453

6. Xavier J, Ward MC, Corr PG, Kalita N, McDonald P. Identifying the barriers and facilitators to culturally responsive HIV and PrEP screening for racial, ethnic, sexual, and gender minoritized patients: A scoping review protocol. PLOS One. doi:10.1371/journal.pone.0281173.

7. Ashana DC, D’Arcangelo N, Gazarian PK, et al. “Don’t Talk to Them About Goals of Care”: Understanding Disparities in Advance Care Planning. J Gerontol A Biol Sci Med Sci. 2022;77(2):339–346. doi:10.1093/gerona/glab091

8. Casanova-Perez R, Apodaca C, Bascom E, et al. Broken down by bias: Healthcare biases experienced by BIPOC and LGBTQ+ patients. AMIA Annu Symp Proc. 2022;2021:275–284.

9. Kabir, R., & Zaidi, S. T. (2022). Implicit bias against BIPOC patients in clinical settings: A qualitative review. Spectra Undergraduate Research Journal. 2022;2(1); 28–46. doi:10.9741/2766-7227.1014

10. Malik S, Master Z, Parker W, DeCoster B, Campo-Engelstein L. In our own words: a qualitative exploration of complex patient-provider interactions in an LGBTQ population. Canadian Journal of Bioethics. 2019;2(2):83–93. doi:10.7202/1062305ar

11. Moore C, Dukes C. The Value of Identity: Providing Culturally-Responsive Care for LGBTQ+ Patients Through Inclusive Language and Practices. Dela J Public Health. 2019;5(3):6–8. doi:10.3281/djph.2019.06.003

12. Minnican C, O’Toole G. Exploring the incidence of culturally responsive communication in Australian healthcare: the first rapid review on this concept. BMC Health Serv Res. 2020;20(1):20. doi:10.1186/s12913-019-4859-6

13. Govere L, Govere EM. How Effective is Cultural Competence Training of Healthcare Providers on Improving Patient Satisfaction of Minority Groups? A Systematic Review of Literature. Worldviews Evid Based Nurs. 2016;13(6):402–410. doi:10.1111/wvn.12176

14. Alizadeh S, Chavan M. Cultural competence dimensions and outcomes: a systematic review of the literature. Health Soc. Care Community. 2015;24(6):e117–e130. doi:10.1111/hsc.12293

15. Morgan KM, Maglalang DD, Monnig MA, Ahluwalia JS, Avila JC, Sokolovsky AW. Medical Mistrust, Perceived Discrimination, and Race: a Longitudinal Analysis of Predictors of COVID-19 Vaccine Hesitancy in US Adults. J Racial Ethn Health Disparities. 2023;10(4):1846–1855. doi:10.1007/s40615-022-01368-6

16. Pharr JR, Terry E, Wade A, Haboush-Deloye A, Marquez E, Nevada Minority Health And Equity Coalition. Impact of COVID-19 on Sexual and Gender Minority Communities: Focus Group Discussions. Int J Environ Res Public Health. 2022;20(1):50. doi:10.3390/ijerph20010050

17. Ndugga N, Hill L, Artiga S. Covid-19 cases and deaths, vaccinations, and treatments by race/ethnicity as of fall 2022. KFF website. November 17, 2022. Accessed May 24, 2023. https://www.kff.org/racial-equity-and-health-policy/issue-brief/covid-19-cases-and-deaths-vaccinations-and-treatments-by-race-ethnicity-as-of-fall-2022/

18. Singu S, Acharya A, Challagundla K, Byrareddy SN. Impact of Social Determinants of Health on the Emerging COVID-19 Pandemic in the United States. Front. Public Health. 2020;8:406. doi:10.3389/fpubh.2020.00406

19. Dey T, Dominici F. COVID-19, Air Pollution, and Racial Inequity: Connecting the Dots. Chem Res Toxicol. 2021;34(3):669–671. doi:10.1021/acs.chemrestox.0c00432

20. Cahill S, Grasso C, Keuroghlian A, Sciortino C, Mayer K. Sexual and Gender Minority Health in the COVID-19 Pandemic: Why Data Collection and Combatting Discrimination Matter Now More Than Ever. Am J Public Health. 2020;110(9):1360–1361. doi:10.2105/AJPH.2020.305829

21. Pharr JR, Batra K. Physical and Mental Disabilities among the Gender-Diverse Population Using the Behavioral Risk Factor Surveillance System, BRFSS (2017-2019): A Propensity-Matched Analysis. Healthcare (Basel). 2021;9(10):1285. doi:10.3390/healthcare9101285

22. Ferlatte O, Salway T, Rice SM, Oliffe JL, Knight R, Ogrodniczuk JS. Inequities in depression within a population of sexual and gender minorities. J Ment Health. 2020;29(5):573–580. doi:10.1080/09638237.2019.1581345

23. Chan ASW, Wu D, Lo IPY, Ho JMC, Yan E. Diversity and Inclusion: Impacts on Psychological Wellbeing Among Lesbian, Gay, Bisexual, Transgender, and Queer Communities. Front Psychol. 2022;13:726343. doi:10.3389/fpsyg.2022.726343

24. Pharr JR, Kachen A, Cross C. Health Disparities Among Sexual Gender Minority Women in the United States: A Population-Based Study. J Community Health. 2019;44(4):721–728. doi:10.1007/s10900-019-00631-y

25. Swann G, Stephens J, Newcomb ME, Whitton SW. Effects of sexual/gender minority- and race-based enacted stigma on mental health and substance use in female assigned at birth sexual minority youth. Cultur Divers Ethnic Minor Psychol. 2020;26(2):239–249. doi:10.1037/cdp0000292

26. Dawson L, McGough M, Kirzinger A, Sparks G, Rae M, Young G, Kates J. The Impact of the COVID-19 Pandemic on LGBT+ People’s Mental Health. KFF website. August 27, 2021. Accessed May 24, 2023. https://www.kff.org/coronavirus-covid-19/poll-finding/the-impact-of-the-covid-19-pandemic-on-lgbt-people/

27. Pharr JR, Terry E, Wade A, Haboush-Deloye A, Marquez E, Nevada Minority Health And Equity Coalition. Impact of COVID-19 on Sexual and Gender Minority Communities: Focus Group Discussions. Int J Environ Res Public Health. 2022;20(1):50. doi:10.3390/ijerph20010050

28. Kriss JL, Hung MC, Srivastav A, et al. COVID-19 Vaccination Coverage, by Race and Ethnicity - National Immunization Survey Adult COVID Module, United States, December 2020-November 2021. MMWR Morb Mortal Wkly Rep. 2022;71(23):757–763. doi:10.15585/mmwr.mm7123a2

29. Ndugga N, Hill L, Artiga S, Haldar S. Covid-19 cases and deaths, vaccinations, and treatments by race/ethnicity as of fall 2022. KFF website. July 14, 2022. Accessed May 24, 2023. https://www.kff.org/coronavirus-covid-19/issue-brief/latest-data-on-covid-19-vaccinations-by-race-ethnicity/

30. Roy DN, Biswas M, Islam E, Azam MS. Potential factors influencing COVID-19 vaccine acceptance and hesitancy: A systematic review. PLoS One. 2022;17(3):e0265496. doi:10.1371/journal.pone.0265496

31. Moore R, Purvis RS, Hallgren E, et al. Motivations to Vaccinate Among Hesitant Adopters of the COVID-19 Vaccine. J Community Health. 2022;47(2):237–245. doi:10.1007/s10900-021-01037-5

32. Jaiswal J, Halkitis PN. Towards a More Inclusive and Dynamic Understanding of Medical Mistrust Informed by Science. Behav Med. 2019;45(2):79–85. doi:10.1080/08964289.2019.1619511

33. Prather C, Fuller TR, Jeffries WL 4th, et al. Racism, African American Women, and Their Sexual and Reproductive Health: A Review of Historical and Contemporary Evidence and Implications for Health Equity. Health Equity. 2018;2(1):249–259. doi:10.1089/heq.2017.0045

34. Logie CH. Lessons learned from HIV can inform our approach to COVID-19 stigma. J Int AIDS Soc. 2020;23(5):e25504. doi:10.1002/jia2.25504

35. Batteux E, Mills F, Jones LF, Symons C, Weston D. The Effectiveness of Interventions for Increasing COVID-19 Vaccine Uptake: A Systematic Review. Vaccines (Basel). 2022;10(3):386. doi:10.3390/vaccines10030386

36. Cascini F, Pantovic A, Al-Ajlouni YA, et al. Social media and attitudes towards a COVID-19 vaccination: A systematic review of the literature. EClinicalMedicine. 2022;48:101454. doi:10.1016/j.eclinm.2022.101454

37. Gibson C, Smith D, Morrison AK. Improving Health Literacy Knowledge, Behaviors, and Confidence with Interactive Training. Health Lit Res Pract. 2022;6(2):e113–e120. doi:10.3928/24748307-20220420-01

38. Schut RA. Racial disparities in provider-patient communication of incidental medical findings. Soc Sci Med. 2021;277:113901. doi:10.1016/j.socscimed.2021.113901

39. Savoia E, Piltch-Loeb R, Goldberg B, et al. Predictors of COVID-19 Vaccine Hesitancy: Socio-Demographics, Co-Morbidity, and Past Experience of Racial Discrimination. Vaccines (Basel). 2021;9(7):767. doi:10.3390/vaccines9070767

40. Arksey H O’Malley L. Scoping studies: Towards a methodological framework. Int J Soc Res Methodol. 2005;8(1):19–32. doi:10.1080/1364557032000119616

41. Munn Z, Peters MDJ, Stern C, Tufanaru C, McArthur A, Aromataris E. Systematic review or scoping review? Guidance for authors when choosing between a systematic or scoping review approach. BMC Med Res Methodol. 2018;18(1):143. doi:10.1186/s12874-018-0611-x

42. Tricco AC, Lillie E, Zarin W, et al. PRISMA Extension for Scoping Reviews (PRISMA-ScR): Checklist and Explanation. Ann Intern Med. 2018;169(7):467–473. doi:10.7326/M18-0850

43. Chaudhary JK, Yadav R, Chaudhary PK, et al. Insights into COVID-19 Vaccine Development Based on Immunogenic Structural Proteins of SARS-CoV-2, Host Immune Responses, and Herd Immunity. Cells. 2021;10(11):2949. doi:10.3390/cells10112949

44. Bell DAA. “Who’s Afraid of Critical Race Theory?” University of Illinois Law Review. 1995;4:893–910.

45. Alexander BK. Queer/Quare Theory. In: Denzin NK, Lincoln YS, eds. The Sage Handbook of Qualitative Research. 5th Edition. SAGE Publishing; 2017:275–307.

46. McLeroy KR, Bibeau D, Steckler A, Glanz K. An ecological perspective on health promotion programs. Health Educ Q. 1988;15(4):351–377. doi:10.1177/109019818801500401

